# The application value of Kano Model in quality of healthcare: A scoping review

**DOI:** 10.1101/2024.03.30.24305047

**Authors:** Xueqing Wang, Panpan Tang, Yueying Jiang, Yuan Zhao, Leiwen Tang, Shina Qiao, Jing Shao, Dandan Chen

## Abstract

**Background:** The Kano model has been utilized in healthcare services with a focus on capturing the patient’s perspective, in order to generate reliable evidence regarding areas that require improvement.

**Objective:** The objective of this study is to provide a description of the implementation of the Kano Model in the healthcare sector and to identify the common quality attributes that have the potential to enhance patients’ satisfaction.

**Methods:** Literature searches were performed using PubMed, Web of Science, Embase, Cochrane, CINAHL, Scopus, OVID, CBM, CNKI, VIP, and Wanfang Database. Primary studies of Kano model and healthcare quality in English and Chinese were included.

**Results:** 42 studies were included eventually. There are three findings: (1) the Kano model’s application is not uniform; (2) dimensions of reliability, efficiency, and assurance are crucial for improving healthcare quality and satisfaction, but tangible and empathy are difficult to interpret; (3) applying Kano model to improve patient satisfaction is imperative.

**Conclusions:** The impact of specific quality attributes on healthcare services and their application in practice are highlighted in this review. Besides, future research could focus on ’Internet+’ nursing applications.

## 1. Introduction

Healthcare quality plays a pivotal role in the medical industry, as it significantly impacts patient satisfaction and treatment outcomes (Shaheen et al., 2019). Kalf (2022) conducted a study which revealed a significant correlation between the quality of care and survival rates among patients diagnosed with breast, lung, and prostate cancers. This finding underscores the importance of prioritizing care quality in the treatment of these conditions. Furthermore, Shie (2022) has suggested that there exists a positive correlation between the health service quality and patient loyalty and satisfaction. Kruk (2018) and the World Health Organization (WHO, 2018) have put forth significant reports emphasizing the need for the healthcare system to prioritize the improvement of care quality and reduce harm, particularly in low- and -middle-income countries. This urgency arises from the growing demand for healthcare services and the limited availability of resources (Zhou et al., 2017). Unfortunately, despite substantial investment and extensive research, the quality of healthcare quality in all countries falls significantly below the desired standards (Berwick et al., 2018). As a consequence of insufficient management of healthcare services and deficiencies in healthcare quality, there have been instances of adverse events in hospitals, excessive and inappropriate care, and other public health concerns that have led to fatalities (Tello et al., 2020).

According to a WHO (2018) report, low-quality health systems countries may contribute to an estimated 8.6 million deaths each year. These deaths could have been prevented if high-quality care had been provided. The National Academies of Sciences (2018) have demonstrated that the issue of the "quality chasm" satisfaction among patients was the availability of pertinent a global scale, even after twenty years of development..

Enhancing patient value and satisfaction represent the primary objective of every healthcare system. To listen to "patients‘ voice", Corbella Jané (2003) introduced Kano Model into the healthcare service industry for the first time in 2003, which was established a "two-dimensional" cognitive system between whether product quality satisfies customers and customer satisfaction as well as considering patients’ emotions and perceptions (Chang & Chang, 2013). The Kano model categorizes quality attributes into five categories: (1) mandatory or "must" (M), (2) Attraction (A), (3) One-dimensional (O), (4) Reverse (R), and (5) Doesn’t matter (I) (Matzler (1996). Most recent studies employ Kano models to identify quality attributes that address patient needs and improve service quality. These studies have demonstrated a reduction in adverse events, improved prognosis, and increased patient (Barrios-Ipenza et al., 2021; Chen et al., 2019; Lin et al., 2022) . However, the healthcare system comprises numerous subsystems, including hospitals with specialty units, clinics, and daycare centers, among others. These subsystems differ in terms of the type of care provided, the services offered, and various other characteristics. The majority of previous research has primarily concentrated on presenting individual findings pertaining to particular specific service units within the healthcare system. However, there is a noticeable dearth of research that offers a comprehensive understanding of patient satisfaction by integrating all service units. Notably, variations exist in the criteria assessed for identical quality attributes (Kuo et al., 2011; Wang et al.,2021), leading to divergent outcomes even when examining the same region (Hejaili et al,. 2009). Consequently, it poses a challenge for a singular study to offer comprehensive guidance for practical application. Therefore, it is imperative to incorporate the typical quality attributes and findings from previous research in order to enhance their systematicity and practicality. This review examines the utilization of the Kano Model in the healthcare service sector and its integration with other quality approaches. It also highlights the significant findings and practices derived from these studies.

## 2. Methods

### 2.1 Protocol and Registration

This review was conducted using Arksey and O’Malley’s methodology framework (Arksey & O’Malley, 2005) and presented in accordance with the PRISMA-ScR (Preferred Reporting Items for Systematic review and Meta-Analysis extension for Scoping Review) checklist (Tricco et al., 2018). This scoping review has been registered with Open Science Framework (https://osf.io/tegmd/).

### 2.2 Eligibility and Criteria

The inclusion criteria for this review were: (1) If the research was a cross-sectional study using the Kano model; (2) if the study was conducted in a health care setting (e.g., hospital, clinic, nursing home, etc.), (3) if the study investigators were nurses or other health care providers, (4) if the study subject was patients or other recipients who received health care service, (5) if the study was an peer reviewed article, and (6) if the study was written in Chinese or English. Exclusion criteria included the following: (1) if they were reviews, letters, conference abstracts, or news, (2) if the full text is not available, (3) if the incomplete report.

### 2.3 Information sources

The following databases were searched from January 1984 to September 2022: PubMed, Web of Science, Embase, The Cochrane Library, CINAHL, Scopus, OVID, China Biomedical Literature Database, China National Knowledge Infrastructure (CNKI), VIP Chinese Science and Technology Journal Database, and Wanfang Database. As Kano Model was proposed in 1984, the starting year for this scoping review was chosen accordingly.

### 2.4 Search

The search terms (keywords) used were Kano Model and healthcare quality. To ensure adequate coverage of the search, it was conducted by MeSH Terms and Entry Terms. Table 1 outlines the search strategy of PubMed used.

**Table 1:**
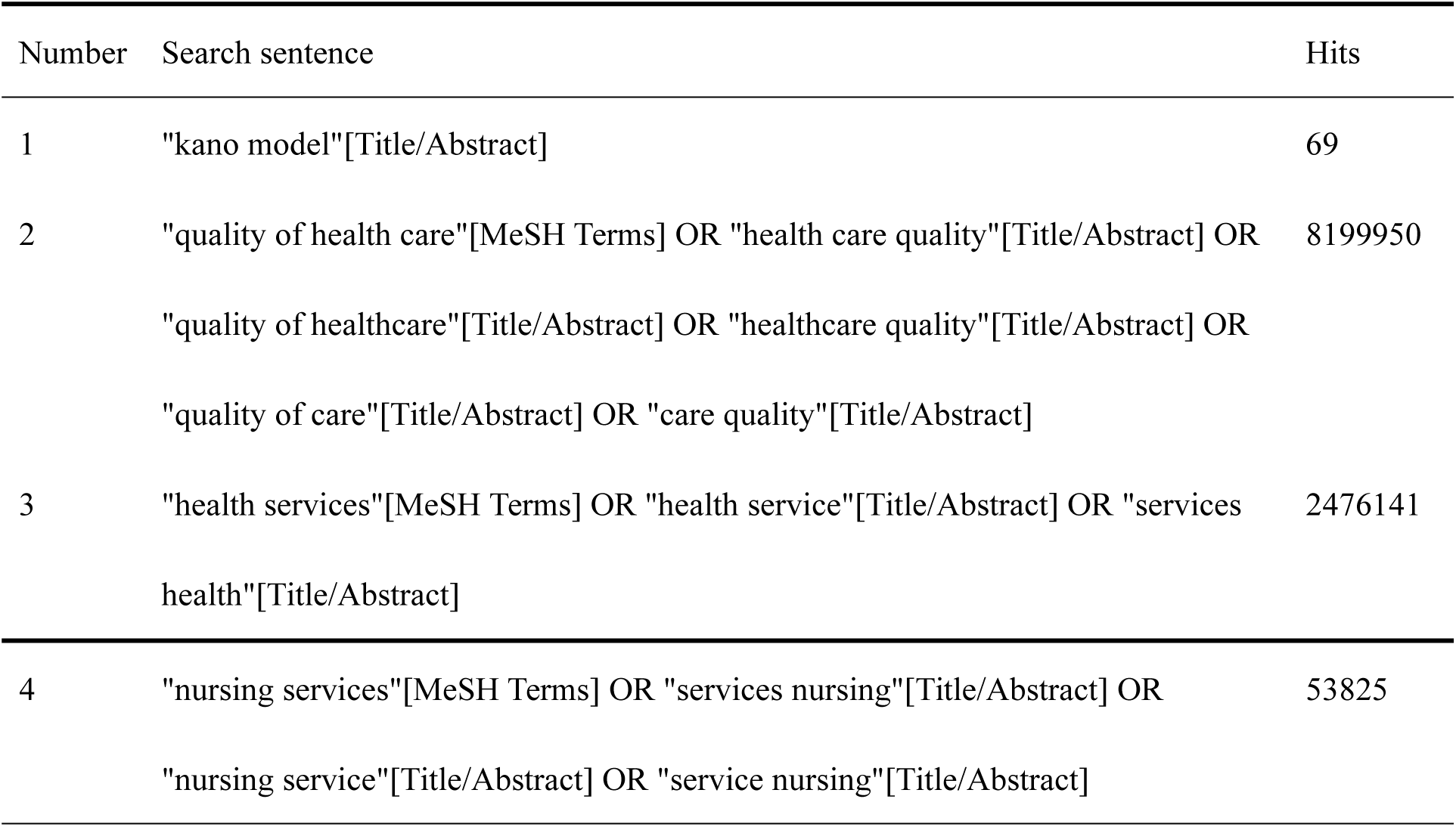
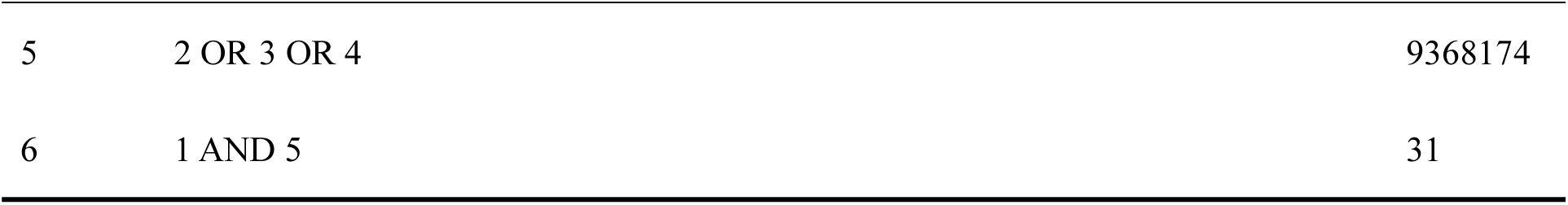
Pubmed Search Strategy.

### 2.5 Selection of evidence

The Zotero software was used to manage the reference and remove duplications. Then two reviewers independently screened the title and abstract of citations to identify articles that meet the minimum inclusion criteria. Disagreements were discussed with a third researcher and resolved by consensus. When a review was found, studies included in the review that met the inclusion criteria were taken into accounted and cross-checked with the list of retrieved articles to ensure no papers had been missed.

### 2.6 Date charting process

To create a descriptive summary of the results, data were charted by extracting: (1) author, year, and country; (2) design; (3) sample;(4) kano categories; (5) usage; (6) intervention; (7) evaluation; and (8) result. Two reviewers independently charted the extracted data from each eligible article and discussed disagreements.

### 2.7 Synthesis result

Reports vary in form, from simply narrating and describing the study’s characteristics to presenting the common quality attributes and applied results. Tables were used to compare the features of the studies and extract data. The original quality attributes were presented in the **supplementary table** (Table S1), extracted directly from the included articles and categorized according to the five categories of Kano Model. Additionally, the original quality attributes (Table S1) were integrated and analyzed in accordance with the five attribute categories of Kano Model. The analysis involved quantifying categories to which each quality attribute in the included article belonged, followed by and then calculating the percentage to ascertain the corresponding category for each attribute. Importantly, the attributes were categorized into five dimensions, namely tangibles, efficiency, reliability, empathy, and assurance.

## 3 Result

### 3.1 Selection of sources of evidence

The initial search identified 1174 articles and removed 689 items after duplicates. Following the screening of titles and abstracts, a total of 80 articles were selected for full-text evaluation. In the end, 42 articles were incorporated into the dataset. (see Fig.1)

**Fig.1.**
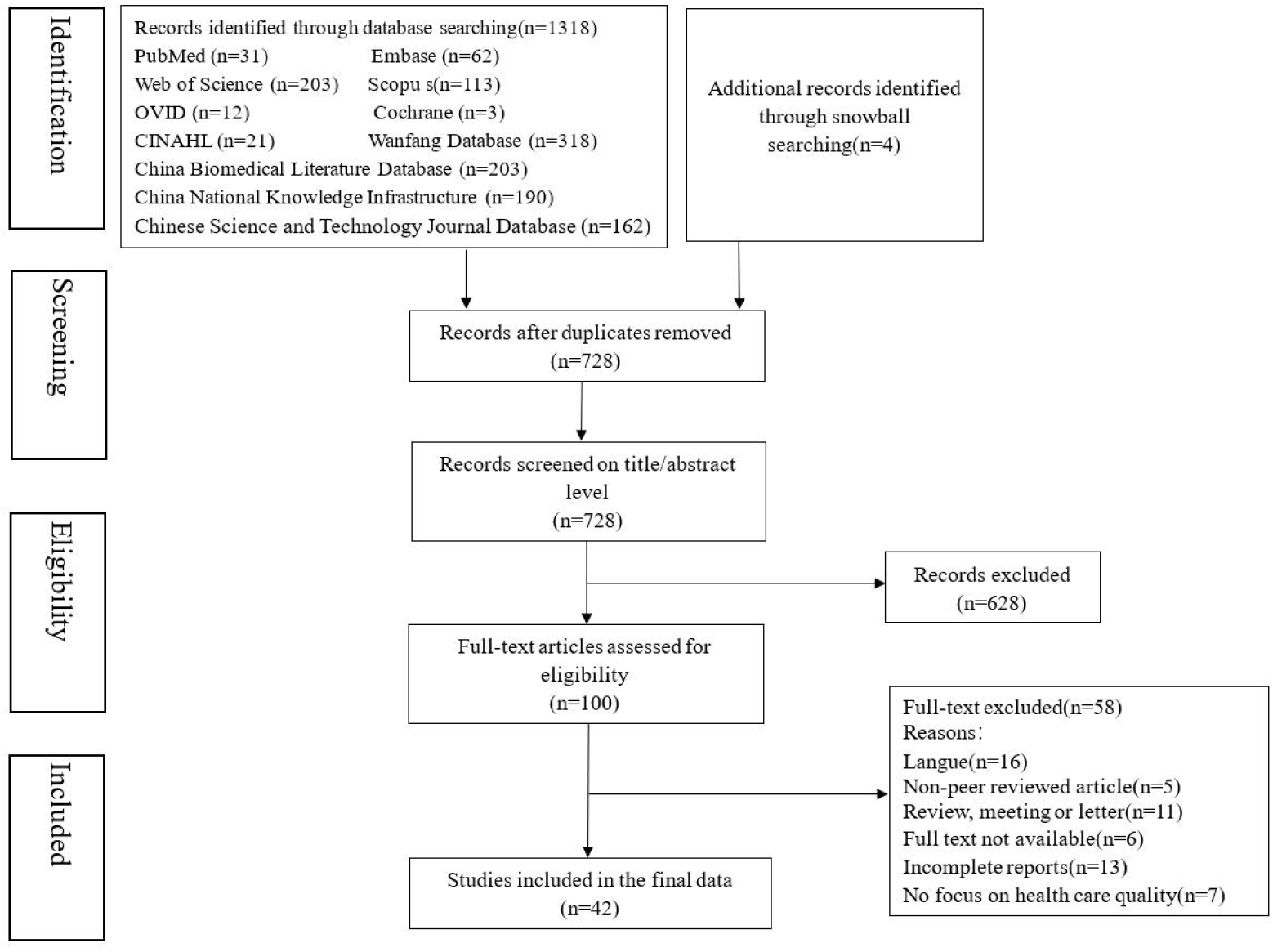
PRISMA flowchart

### 3.2 Characteristics of evidence

The selected studies were published between 2005 and 2023. A total of 33 (76.2%) were published in or after 2015. The most studies (n=21) were conducted in inpatient department, following was outpatient department (n=11), emergency department (n=4), basic health unites (n=2), nursing home (n=1), walk-in clinic (n=1), school infirmary (n=1), and public hospital (n=1). Study designs included 32 cross-section studies and 10 contained both cross-sectional and experimental studies. The number of quality attributes varies from 4 to 50. Thirty-two studies used only Kano Model, five studies integrated Kano Model with the SERVQUAL Model, and three articles mixed Kano Model integrated the Quality Function Development (QFD).

### 3.3 Synthesis of results

#### 3.3.1 The common quality attributes

From all the reports, we summarized diverse quality attributes (n=63) into five dimensions: (1) Tangibles, (2) Efficiency, (3) Reliability, (4) Empathy, and (5) Assurance, which are borrowed from the SERVQUAL model. Further descriptions of these quality attributes are reported in Table 3.

##### 3.3.1.1 Tangible

This dimension pertains to the appearance of physical facilities, equipment, personnel, and communication materials. Sixteen quality attributes are encompassed within this dimension. Among these attributes, three fall under the must-be category, indicating that patients will experience significant dissatisfaction if these services fail to meet their needs. Fifteen studies (Chiou & Cheng, 2008; Ke et al., 2021; Patel & Bhatt, 2017; He Jian-qun et al., 2022; Yao Yao et al., 2008; Zhang Zhuo-yi et al., 2013; Li Zhen, 2017; Su Yin, 2013; LI Teng-hun et al., 2015; Chen et al., 2020; Xiong Wei et al., 2012; Xiong et al., 2022; Wang Huaiyan et al., 2021; Nie Shijun, 2016;Gong et al., 2019) found patients place significant importance on the treatment environment, considering a "comfortable and hygienic hospital environment" as a fundamental requirement.

The one-dimensional category comprises seven quality attributes, with the highest proportion being “maternal and child room.“ Li Zhen (2017), Su Yin (2013), Wang Huaiyan (2021), and Gong (2019) have suggested that setting up maternal and child rooms can help mitigate disputes, complaints. Six researchers have identified three quality attributes that fall under the attractive category, with none being classified as indifferent or reverse. On the other hand, there are two attributes that do not effectively differentiate distinguish the category, namely "accessible educational brochures" and "colorful wall".

##### 3.3.1.2 Efficiency

In the context of healthcare system, efficiency pertains to the effective utilization of resources and the management of waiting time in processes. As shown in Table 3, this dimension comprises eleven quality attributes. The majority of them (n=8) are categorized as the one-dimensional attributes, indicating that individuals have certain expectations for these services and their direct impact on patient satisfaction.

According to twelve studies (Barrios-Ipenza et al., 2021; Ferreira et al., 2021; He et al., 2020; LI Teng-hun et al., 2015; Chen et al., 2022; Hu et al., 2011; He Jian-qun et al., 2022; Tao Yao et al., 2008; Zhang Qunxiang et al., 2021; Xiong Wei et al., 2012; Hun Chen et al., 2020), "the reasonable waiting time" is considered a mutual agreement between the healthcare system and patients, with the potential to greatly enhance patient satisfaction. In contrast, patients may perceive a prolonged waiting time as a breach of promise (Godley & Jenkins, 2019). Only one study involved "the level of bureaucracy" and regarded it as a must-be quality. None of the quality attributes belong to the attractive, indifferent, or reverse category. Meanwhile, the categorization of "timely resolution of potential problems" and "easy retrieval of examination results" be determined.

##### 3.3.1.3 Reliability

Sixteen quality attributes are used to assess the dependability and accuracy of the promised service, as depicted in Table 3. Eight quality attributes belong to the one-dimensional. The two most prominent factors pertaining to security are "reliability and assurance of information" and "expected therapeutic effect," constituting a substantial proportion. Kuo (2011) demonstrated that security plays a crucial role in supporting healthcare services and can effectively minimize the burden on resources and time.

Six attributes fall into the must-be category, with " professional skills" being the attribute most commonly cited. None attribute belongs to the attractive, the category and the reverse category. Moreover, it is challenging to determine the category to which certain attributes (n=2) belong.

##### 3.3.1.4 Empathy

This dimension pertains to the delivery of personalized and attentive care to customers. Among the one-dimensional category, there are five quality attributes, with "attention to patient needs" being the most frequently cited attribute. Xiong et al. (2022) propose that nurses should adopt a proactive approach by actively inquiring about patients’ needs in order to enhance compliance and enhance overall satisfaction. Three quality attributes belong to the attractive category. "Proper health education" is the most frequently cited factor, indicating that patients have a pressing demand for health education (Tian Yuan et al., 2017).

"Adequate protecting patients’ privacy" and "comfortable for invasive operations" belongs to the must-be category, and "respectful of religious belief" is viewed as an indifferent attribute. On the other hand, the categorization of "resolution of the complaint" remains undetermined..

##### 3.3.1.5 Assurance

This dimension encompasses eight quality attributes that exemplify employees’ knowledge, courtesy, and ability to convey trust and confidence. Five quality attributes lie in the one-dimensional category. Among them, "kindness and courteous of personnel" merges as the most frequent attributes.

Barrios-Ipenza (2021) study illustrated that patient satisfaction is likely to increase in direct proportion to the presence of "kind and courtesy". "Protection of patients’ legal rights" falls under the category of must-be, as it plays a crucial role in effectively mitigating medical disputes ( He et al., 2020; Zhang Zhuo-yi et al., 2013; Li Zhen, 2017; Su Yin, 2013; Luming Li et al., 2022; Nie Shijun, 2016; Chen et al., 2022). In addition, it is challenging to address two quality attributes within a specific category.

#### 3.3.2 The application of Kano Model

According to the findings of the review, 32 studies exclusively utilized the Kano Model, while 10 papers employ not only Kano Model but also other models for quality improvement. Among them, 5 studies integrated Kano Model with the SERVQUAL Model (C. M. Chou et al., 2015; Gomez Martin et al., 2019; Lacerda et al., 2021; Mehrabian et al., 2021; Patel & Bhatt, 2017). In addition, three articles combined Kano Model with QFD (Chiou & Cheng, 2008; Kuo et al., 2011; Zhang Qunxiang et al., 2021). One study conducted by de Vasconcelos et al. (2022) incorporated the use of the Balanced Scorecard (BSC) as a research tool. Finally, one study employed the Customer Satisfaction Index Model(Hu et al., 2011).

After conducting a thorough analysis of articles that exclusively utilize the Kano Model, it has been observed that the app2013cation of the Kano Model follows a consistent pattern, which can be divided into three distinct stages. Step 1 involves the development of the Kano questionnaire. 9 articles developed it by adapting the existing questionnaire (Ke et al., 2021; Lee et al., 2007; Li Zhen, 2017; Li Teng-hun et al., 2015; Luming Li et al., 2022; Xiong Wei et al., 2012; Wang Shu-yi, 2005; Zhai et al., 2015; Nie Shijun, 2016), and 3 article adopt the method of review articles (Materla & Cudney, 2020; Rozaq et al., 2019; Xiong et al., 2022). Additionally, 4 studies tested the reliability and validity of the questionnaire (Materla & Cudney, 2020; Li, 2021; Luming Li et al., 2022; Zhai et al., 2015). The Kano questionnaire consists of a set of attribute-specific questions. Each attribute is assessed through a pair of questions, comprising functional and dysfunctional aspects (Lee et al., 2007). Step 2 involves the dissemination of the Kano survey, while step 3 entails the analysis of the collected data. At this stage, the categorization of each attribute is determined based on its frequency.

Of the 5 studies using Kano Model and the SERVQUAL Model, 3 papers (Gomez Martin et al., 2019; Mehrabian et al., 2021; Patel & Bhatt, 2017) first used the SERVQUAL Model to determine the gap between patients’ perceptions and expectations of healthcare quality, and then applied Kano Model to identify attributes that significantly influence patients’ satisfaction. Chou (2015) administered both the SERVQUAL questionnaire and Kano questionnaire simultaneously, subsequently identifying the priority service through data analysis. Another recent study utilized the five dimensions of the SERVQUAL Model to develop the questionnaire as Kano Model lacked predefined quality classifications (Lacerda et al., 2021).

Three papers integrated Kano Model and QFD to improve healthcare service quality (Chiou & Cheng, 2008; Kuo et al., 2011; Zhang Qunxiang et al., 2021). Firstly, Kano Model was employed to ascertain the service requirements of patients. Secondly, the Quality Function Deployment (QFD) methodology was utilized to convert these identified quality attributes into a comprehensive service planning framework. The final two models have the potential to compensate for the limitations of the Kano Model. de Vasconcelos (2022) used Kano -BSC to develop a strategic plan for public health. Based on the results of Kano questionnaire, BSC was employed to ascertain the which characteristics have the greatest impact on patient satisfaction. This method can not only analyze quality attributes but also control the cost. Due to the limitations of the Kano Model in analyzing patients’ loyalty and complaints, Hu (2011) combined Kano Model with the Customer Satisfaction Index Model to address this issue.

### 3.3 The effects of using the Kano Model

Nine articles encompass experimental studies (He et al., 2020; Zhang Qi et al., 2022;Chen et al., 2022;Li Zhen, 2017;Su Yin, 2013; Wang Huaiyan et al., 2021; Tian Yuan et al., 2017; Nie Shijun, 2016; Gong et al., 2019). All implemented interventions were derived from the findings of Kano Model. Seven studies were controlled experimental studies (He et al., 2020; Zhang Qi et al., 2022;Chen et al., 2022;Su Yin, 2013; Wang Huaiyan et al., 2021; Tian Yuan et al., 2017; Nie Shijun, 2016), while two studies utilized a before-after design within the same patient population (Li Zhen, 2017; Gong et al., 2019). The study sample sizes exhibited a wide range, varying from 45 to 13689. The primary criterion for evaluation was the patient’s satisfaction(n=9). Other indicators of outcomes included the quality of healthcare service (n=4) and the patient’s emotions (n=2). All nine studies’ results indicated that the implementation of the Kano Model has a positive impact on patient satisfaction rates and enhances the overall quality of healthcare services. Zhang Qi (2022) and Wang Huaiyan (2021) have suggested that Kano Model could significantly improve patients’ emotions, as evidenced by SAS, SDS, and CMFS scores.

## 4 Discussion

### 4.1 Quality attributes of Kano Model

Despite the fact that the Kano Model has been the subject of research for more than four decades and has been extensively applied in healthcare services since 2015, its implementation is still in its early stages. There is a need for enhanced clarity in quality attributes pertaining to healthcare. This review reveals that the most quality attributes fall under the one-dimensional category, with the must-be and attractive categories following closely behind. According to the gathered information, the one-dimensional category was the main characteristic among the five dimensions evaluated. (Kametani et al., 2010). Therefore, in order to increase in patient satisfaction, it is advisable to prioritize the enhancement of one-dimensional attributes. In the field of medical research, it has been established that waiting time a crucial factor in patient (Du et al., 2018). Therefore, it is imperative to have adequate facilities in place. This is observation aligns with the information provided in Table 3, which categorizes "proper and adequate medical equipment" as a one-dimensional attribute. Similarly, the importance of "kindness and courteous service attitude" in influencing patient satisfaction has been emphasized in previous studies (Fang et al., 2019; Yusefi et al., 2022). To enhance the perceived value of healthcare services, it is important to address not only the basic needs and performance satisfaction of patients, but also their emotional needs. According to Nilsson-Witell and Fundin (2005), patients seek attractive quality and unexpected. The study found a negative association between spiritual encouragement received and disability, with the strongest relationship observed among individuals with lower levels of interest in encouragement. Also, support received had a large positive association with satisfaction (McWilliams et al., 2017). The need to pay attention is, compared to the other categories, attributes of the must-be category would generate less satisfaction than those involved in providing services. Still, they are the foundation of the whole healthcare system (Paraschivescu & COTÎRLEŢ, 2012).

From the perspective of dimensions, efficiency, assurance, and reliability were dimensions with a high percentage of one-dimensional quality attributes (over 50%), this results indicate that these quality attribute result in satisfaction when fulfilled and result in dissatisfaction when not fulfilled (Patel & Bhatt, 2017). They are clear and are something the hospital can focus on, so they are both necessary and primary conditions for patient satisfaction. In addition, nine quality attributes in the tangible dimension cannot be clearly categorized, meaning these services need further exploration. The five dimensions showed an insignificant percentage of the indifferent and reverse attributes. According to the dynamic principle of the Kano Model (Nilsson-Witell & Fundin, 2005), the quality attribute is not invariable.

With time, one quality attribute can evolve into other quality attribute, showing the evolution process from indifferent, attractive, one-dimensional, and must-be attributes. Thus, no matter which category the quality attributes belong to, they still need attention.

Due to the shortage of nurses in China, where the nurse-patient ratio is 1:3.56, and the rising costs, nurse managers need to accurately control costs. The Kano model can be used to provide optimal service to patients by determining their needs for healthcare services and meeting their necessary needs. Also, it can integrate patients into the decision, and allocate finite resources reasonably. For example, religious consideration is the indifference, especially in China. Therefore, nurses can pay little attention to it and focus on professional skills or service attitude. On the other hand, the provision of healthcare services that respect and meet the needs of patients is essential to promote positive care outcomes. However, family overprotection can cause nurse-patient disputes and hinder normal healthcare activities. Therefore, nurse managers can significantly improve patient satisfaction by refining the nursing process based on the results of the Kano model.

### 4.2 Kano Model applications and applicable outcomes

The healthcare system has many subsystems, such as outpatient clinics, specialized wards in hospitals, clinics, and daycare centers, which vary according to the type of care, type of service, and various other characteristics (Materla, Cudney, & Antony, 2019). In this review, we find Kano Model has been applied for all most every healthcare setting. It is most commonly used in the investigation of inpatient wards, as shown in Table 2. This might relate to the special case of hospitalized patients, such as fall risk (Choi et al., 2021) and mental health (Yu et al., 2019), which are close to patient satisfaction (Crosier et al., 2012). Outpatient satisfaction is slightly different from inpatient satisfaction (Lv et al., 2016; Rho et al., 2013); they are both connected with the ability of personal healthcare communication, while the waiting time for medical exams affects outpatient satisfaction as well. Therefore, it is necessary to distinguish strategies for improving patient satisfaction among different patient types.

**Table 2:** Characteristics of included studies.

| Author, year, country | Design | Settings of Participants | Number of Quality attributes | Categories of Quality Attribute |  |  |  |  | Model application |
| --- | --- | --- | --- | --- | --- | --- | --- | --- | --- |
|  |  |  |  | M | O | A | I | R |  |
| Wang Shu-yi et al. 2005/China | Descriptive research | Outpatient clinic | 40 | 16 | 17 | 3 | 4 | 0 | Kano Model |
| Lee, Chang, and Chao 2007/Taiwan, China | Descriptive research | Inpatient department | 26 | 0 | 22 | 0 | 4 | 0 | Kano Model |
| Chiou and Cheng 2008/Taiwan, China | Descriptive research | Inpatient department | 4 | 2 | 1 | 1 | 0 | 0 | Kano Model + Quality Function Development (QFD) |
| Yao Yao et al. 2008/China | Descriptive research | Inpatient department | 24 | 3 | 16 | 5 | 0 | 0 | Kano Model |
| Hu et al. 2011/ Taiwan, China | Descriptive research | Inpatient department | 37 | 8 | 26 | 3 | 0 | 0 | Kano Model +the Customer Satisfaction Index Model |
| Kuo et al. 2011/Taiwan,China | Descriptive research | Discharged department | 22 | 0 | 20 | 1 | 1 | 0 | Kano Model + Quality Function Development (QFD) |
| Cordero-Ampuero et al., 2012/ Spanish | Descriptive research | Plastic Surgery Ward | 13 | 0 | 1 | 9 | 2 | 1 | Kano Model |
| Xiong Wei et al. 2012/China | Descriptive research | Outpatient clinic | 20 | 7 | 9 | 3 | 1 | 0 | Kano Model |
| Zhang Zhuo-yi et al. 2013/China | Descriptive research | Emergency department | 50 | 25 | 12 | 8 | 2 | 3 | Kano Model |
| Su Yin 2013/China | Descriptive + Experimental research | Pediatric outpatient and emergency department | 25 | 10 | 9 | 6 | 0 | 0 | Kano Model |
| Chou et al. 2015/Taiwan, China | Descriptive research | Nursing home | 24 | 0 | 6 | 14 | 4 | 0 | Kano Model +SERVQUAL model |
| Li Teng-hun et al. 2015/China | Descriptive research | Outpatient clinic | 21 | 8 | 8 | 3 | 2 | 0 | Kano Model |
| Zhai et al. 2015/China | Descriptive research | Midwife outpatient clinic | 24 | 1 | 10 | 13 | 0 | 0 | Kano Model |
|  |  |  |  | M | O | A | I | R |  |
| Li Zhen 2017/China | Descriptive + Experimental research | Pediatric Emergency department | 23 | 14 | 3 | 3 | 3 | 0 | Kano Model |
| Patel and Bhatt 2017/India | Descriptive research | Inpatient department | 50 | 13 | 34 | 3 | 0 | 0 | Kano Model +SERVQUAL models |
| Tian Yuan et al. 2017/China | Descriptive + Experimental research | Geriatric department | 30 | 5 | 7 | 17 | 1 | 0 | Kano Model |
| Gomez Martin et al. 2019/Spanish | Descriptive quantitatively research | Burn Unit | 11 | 2 | 0 | 3 | 4 | 2 | Kano Model +SERVQUAL model |
| Gong et al. 2019/China | Descriptive + Experimental research | Pediatric ward | 23 | 9 | 9 | 5 | 0 | 0 | Kano Model |
| Rozaq et al. 2019/Indonesia | Descriptive research | Inpatient department | 20 | 0 | 11 | 0 | 9 | 0 | Kano Model |
| Howsawi et al. 2020/Saudi Arabia | Descriptive research | Primary healthcare centers | 16 | 0 | 12 | 3 | 1 | 0 | Kano Model |
| Hun Chen et al. 2020/China | Descriptive research | Inpatient department | 27 | 9 | 13 | 5 | 0 | 0 | Kano Model |
| He et al. 2020/China | Descriptive + Experimental research | Outpatient clinic | 15 | 9 | 3 | 3 | 0 | 0 | Kano Model |
| Tejaswi Materla and Cudney 2020/the USA | Descriptive research | Walk-in clinics | 18 | 1 | 15 | 2 | 0 | 0 | Kano Model |
| Barrios-Ipenza et al. 2021/Peru | Descriptive research | Outpatient clinic | 31 | 3 | 27 | 0 | 0 | 1 | Kano Model |
| Ke, Chen, and Gou 2021/China | Descriptive research | Outpatient clinic | 17 | 9 | 0 | 6 | 2 | 0 | Kano Model |
| Lacerda et al. 2021/Brazil | Descriptive research | Basic Health Units | 23 | 0 | 21 | 1 | 1 | 0 | Kano Model +SERVQUAL models |
| Mehrabian 2021/Iran | Descriptive research | Inpatient department | 25 | 0 | 11 | 13 | 0 | 1 | Kano Model +SERVQUAL models |

| Author, year, country | Design | Settings of Participants | Number of<br>Quality attributes | Categories of Quality Attribute |  |  |  |  | Model application |
| --- | --- | --- | --- | --- | --- | --- | --- | --- | --- |
|  |  |  |  | M | O | A | I | R |  |
| Ferreira et al. 2021/<br>Portugal | Descriptive research | Pediatric ward | 31 | 3 | 27 | 0 | 0 | 1 | Kano Model |
| Wang Huaiyan et al.<br>2021/China | Descriptive +<br>Experimental research | Pediatric outpatient clinic | 15 | 3 | 5 | 4 | 2 | 1 | Kano Model |
| Zhang Qunxiang et<br>al. 2021/China | Descriptive research | Inpatient department | 24 | 2 | 11 | 6 | 5 | 0 | Kano Model +Quality Function<br>Development (QFD) |
| de Vasconcelos et al.<br>2022/Brazil | Descriptive research | Public hospital | 20 | 4 | 16 | 0 | 0 | 0 | Kano Model +the Balanced<br>Scorecard |
| He Jian-qun et al.<br>2022/China | Descriptive research | Physical examination<br>center | 21 | 8 | 9 | 3 | 1 | 0 | Kano Model |
| Chen Chen et<br>al.2022/ China | Descriptive +<br>Experimental research | Outpatient clinic | 25 | 14 | 8 | 0 | 3 | 0 | Kano Model |
| Luming Li et al.<br>2022/China | Descriptive research | Inpatient department | 26 | 5 | 10 | 10 | 1 | 0 | Kano Model |
| Xiong et al.<br>2022/China | Descriptive research | School infirmary | 12 | 3 | 3 | 6 | 0 | 0 | Kano Model |
| Zhang Qi et al.<br>2022/China | Descriptive +<br>Experimental research | Thoracic surgery<br>department | 17 | 9 | 7 | 1 | 0 | 0 | Kano Model |
| Cudney et al., 2023/<br>the USA | Descriptive research | The wound care/ostomy<br>clinic | 17 | 0 | 16 | 1 | 0 | 0 | Kano Model |
| Zhu et al., 2023/<br>China | Descriptive +<br>Experimental research | Pediatric Medical Ward | 25 | 11 | 10 | 4 | 0 | 0 | Kano Model |
| Wang et al., 2023/<br>China | Descriptive research | Cardiology ward | 12 | 2 | 5 | 3 | 1 | 0 | Kano Model |
| Yao Yao et al.,<br>2023/China | Descriptive research | Obstetrical department | 25 | 9 | 8 | 8 | 0 | 0 | Kano Model |
| Sun Weigeet al.,<br>2023/ China | Descriptive research | Breast surgery ward | 27 | 2 | 22 | 2 | 1 | 0 | Kano Model |

M=Must-be, O=One-dimensional, A=Attractive, I=Indifferent, R=Reverse

**Table 3:** The typical quality attributes.

| Dimension | Attribute | Kano Categories |  |  |  |  |
| --- | --- | --- | --- | --- | --- | --- |
|  |  | M | O | A | I | R |
| Tangible | Systematic layout of Hospital departments | 0 | 4 (80%) | 1 (20%) | 0 | 0 |
|  | Proper safety and comfort measures | 6 (23.1%) | 6 (23.1%) | 9 (34.6%) | 5 (19.2%) | 0 |
|  | Proper medical equipment | 1 (6.7%) | 12 (80%) | 2 (13.3%) | 0 | 0 |
|  | Comfortable and hygienic hospital environment | 15 (46.9%) | 8 (25%) | 7 (21.9%) | 2 (6.25%) | 0 |
|  | Nutritious and appropriate meals | 5 (41.7%) | 3 (25%) | 4 (33.3%) | 0 | 0 |
|  | Comfortable single room | 0 | 1 (25%) | 2 (50%) | 1 (25%) | 0 |
|  | Informatization platform | 0 | 7 (58.3%) | 5 (41.7%) | 0 | 0 |
|  | Clear sign | 7 (70%) | 1 (10%) | 2 (20%) | 0 | 0 |
|  | Maternal and child room | 0 | 4 (100%) | 0 | 0 | 0 |
|  | Moving line | 0 | 1 (100%) | 0 | 0 | 0 |
|  | Clean employee's clothing | 1 (12.5%) | 6 (75%) | 0 | 1 (12.5%) | 0 |
|  | Adequate and intact supply (e.g.: water, electricity et.al.) | 4 (36.4%) | 7 (63.6%) | 0 | 0 | 0 |
|  | Accessible educational brochures | 2 (28.6%) | 2 (28.6%) | 2 (28.6%) | 1 (14.3%) | 0 |
|  | Voice interactive intelligent robot | 0 | 0 | 1 (100%) | 0 | 0 |
|  | Set up a complaint mailbox | 0 | 0 | 0 | 1 (100%) | 0 |
|  | Colorful wall | 0 | 0 | 1 (50%) | 1 (50%) | 0 |
| Efficiency | The level of bureaucracy | 1 (100%) | 0 | 0 | 0 | 0 |
|  | Reasonable waiting time for a medical consultation | 1 (6.25%) | 12 (75%) | 3 (18.75%) | 0 | 0 |
|  | Time spent on patient care | 1 (33.3%) | 2 (66.7%) | 0 | 0 | 0 |
|  | Business hours | 1 (9.1%) | 8 (72.7%) | 2 (18.2%) | 0 | 0 |
|  | Convenience of transportation | 3 (21.4%) | 9 (64.2%) | 1 (7.1%) | 1 (7.1%) | 0 |
|  | Timeliness of disease diagnosis | 1 (25%) | 3 (75%) | 0 | 0 | 0 |
|  | Easy and convenient registration | 1 (14.3%) | 6 (85.7%) | 0 | 0 | 0 |
|  | Timely resolution of potential problems | 1 (33.3%) | 1 (33.3%) | 1 (33.3%) | 0 | 0 |
|  | Easy retrieval of examination results | 0 | 2 (50%) | 2(50%) | 0 | 0 |
|  | High efficiency of hospitalization procedures | 2 (15.4%) | 9 (69.2%) | 2 (15.4%) | 0 | 0 |
|  | Reasonable arrangement of physical examination | 2 (33.3%) | 3(50%) | 1 (16.7%) | 0 | 0 |
|  |  | M | O | A | I | R |
| Reliability | Professional skills | 21 (52.5%) | 14 (35%) | 4 (10%) | 1(2.5%) | 0 |
|  | Detailed explanation for diagnosis and treatment | 13 (43.3%) | 11 (36.7%) | 6 (20%) | 0 | 0 |
|  | Quick staff response | 6 (42.9%) | 5 (35.7%) | 3 (21.4%) | 0 | 0 |
|  | Timely provide the promised service on | 0 | 9 (56.3%) | 7 (43.7%) | 0 | 0 |
|  | Regular medications | 1 (20%) | 3 (60%) | 0 | 1 (20%) | 0 |
|  | Well-arranged medical rounds and treatment | 2 (28.6%) | 3 (42.8%) | 2 (28.6%) | 0 | 0 |
|  | Well-scheduled patient activities | 1 (25%) | 2 (50%) | 0 | 1 (25%) | 0 |
|  | All equipment work properly | 0 | 2(66.7%) | 1 (33.3%) | 0 | 0 |
|  | Good quality medicines | 1 (50%) | 1 (50%) | 0 | 0 | 0 |
|  | Expected therapeutic effect | 5 (29.4%) | 12 (70.6%) | 0 | 1 | 0 |
|  | No side effect | 6 (66.7%) | 1 (11.1%) | 1 (11.1%) | 0 | 1 (11.1%) |
|  | Reliability and assurance of information | 2 (16.7%) | 10 (83.3%) | 2 (16.7%) | 0 | 0 |
|  | Continuous whole-course team service | 0 | 1 (50%) | 1 (50%) | 0 | 0 |
|  | Active rescue | 0 | 1(100%) | 0 | 0 | 0 |
|  | Correct and completed case history | 2 (50%) | 1 (25%) | 1 (25%) | 0 | 0 |
|  | Accurate bills | 12 (42.9%) | 11 (39.3%) | 2 (7.1%) | 2 (7.1%) | 1 (3.6%) |
| Empathy | Personalized care | 2 (22.2%) | 6 (66.7%) | 0 | 1(11.1%) | 0 |
|  | Attention to patient needs | 2 (6.7%) | 22 (73.3%) | 4 (13.3%) | 2 (6.7%) | 0 |
|  | Resolution of complaints | 2 (40%) | 1 (20%) | 2(40%) | 0 | 0 |
|  | Good communication and patient engagement | 6 (22.2%) | 18 (66.7%) | 2 (7.4%) | 1 (3.7%) | 0 |
|  | No discrimination to the patients | 1 (25%) | 3 (75%) | 0 | 0 | 0 |
|  | Psychological treatment for relatives | 1 (16.7%) | 0 | 4 (66.7%) | 1(16.7%) | 0 |
|  | Priority to patient' interests | 1 (25%) | 1(25%) | 2 (50%) | 0 | 0 |
|  | Adequate protection of patient's privacy | 7 (46.7%) | 5 (41.7%) | 2 (16.7%) | 1 (8.3%) | 0 |
|  | Respectful of religious belief | 0 | 1 (33.3) | 0 | 2 (66.7%) | 0 |
|  | Emotional comfort | 0 | 2(100%) | 0 | 0 | 0 |
|  | Comfortable for invasive operations | 1(100%) | 0 | 0 | 0 | 0 |
|  | Proper health education | 9 (20.5%) | 14 (31.8%) | 17 (38.6%) | 4(9.1%) | 0 |
|  |  | M | O | A | I | R |
| Assurance | Kindness and courteous service attitude | 14 (30.4%) | 25 (54.3%) | 6 (13.1%) | 1(2.2%) | 0 |
|  | Feeling safe at the hospital | 0 | 3 (50%) | 1(25%) | 1(25%) | 0 |
|  | Spiritual encouragement | 1 (7.1%) | 9 (64.3%) | 4 (28.6%) | 0 | 0 |
|  | Sympathetic staff | 0 | 2(100%) | 0 | 0 | 0 |
|  | Insurance consultant | 0 | 1 (50%) | 1(50%) | 0 | 0 |
|  | Understandable expression | 1 (50%) | 0 | 0 | 1 (50%) | 0 |
|  | Protection of patients' legal rights | 7 (46.7%) | 5 (33.3%) | 3 (20%) | 0 | 0 |
|  | Good medical ethics | 0 | 5 (100%) | 0 | 0 | 0 |

M=Must-be, O=One-dimensional, A=Attractive, I=Indifferent, R=Reverse

**Table 4:** the applicable results of Kano Model.

| Author, year, country | Design | Sample size | Intervention | Evaluation | Result |
| --- | --- | --- | --- | --- | --- |
| Su Yin<br>2013/China | Controlled<br>experimental study | 292 | Strengthen the professional training for nurses;<br>Improve the medical environment;<br>Strengthen health education | The rate of patient's satisfaction | Increase satisfaction ratio |
| Nie Shijun<br>2016/China | Controlled<br>experimental study | 125 | Improve hardware equipment;<br>Improve nurses' professional knowledge and emergency response ability;<br>Strengthen communication ability;<br>Enhance health education;<br>improve basic daily necessities | Satisfaction of children's families;<br>Quality of care | ① Increase satisfaction ratio<br>② Improved the scores of nursing service quality in all aspects |
| Tian Yuan et al.<br>2017/China | Controlled<br>experimental study | 186/190 | Practice must be quality attributes;<br>Provide rehabilitation function exercise guidance;<br>Detailed discharge guidance;<br>Explanation of patients' questions; Provide health education | Satisfaction of healthcare service | The satisfaction of the observation group was significantly higher than the control group |
| Li Zhen<br>2017/China | before-after study<br>in the same patient | 285 | Improve the environment;<br>Improve professional skills<br>Develop a good medical attitude; Increase health education | The rate of patient's satisfaction | Increase satisfaction ratio |
| Gong et al.<br>2019/China | before-after study<br>in the same patient | 227 | Strengthen the ability to quickly and accurately identify critically ill children;<br>Improve the inspection system;<br>Improve various operation standards;<br>Pay attention to health education;<br>Improve the ward environment | Satisfaction of children's families;<br>Quality of care | ① The scores of treatment environment, service attitude, operation technology and health education rise<br>② Reduced the rates of disputes and complaints |
| He et al.<br>2020/China | Controlled<br>experimental study | 13689 | Enhance professional skills and etiquette | The rate of patient's satisfaction;<br>The defect rate in outpatient clinic | ① Increase satisfaction ratio;<br>② Reduce the number of clinic defects |
| Wang<br>Huaiyan et al.<br>2021/China | Controlled<br>experimental study | 45 | Improve medical environment, service attitude and professional skills | Nursing satisfaction of parents;<br>Nursing quality score;<br>Fear during the visit;<br>Visit compliance | ① Healthcare service satisfaction in the observation group was higher than control group;<br>② The scores of medical environment, service attitude, professional skills and health education in the observation group were higher than the control group;<br>③ Frankl scale score in the observation group was higher than the control group<br>④ CMFS score was lower than the control group |
| Chen et al.2022/<br>China | Controlled<br>experimental study | 329 | Improve the medical environment<br>Convenient consultation and self-service;<br>Improve the construction of specialized nursing clinics;<br>Examination report interpretation service;<br>Medication guidance | The rate of patient's satisfaction | Increased patient satisfaction |
| Zhang Qi et al.<br>2022/China | Controlled<br>experimental study | 120 | Practice must be quality attributes; Provide rehabilitation function exercise guidance;<br>Discharge guidance;<br>Provide health education | Health knowledge mastery;<br>Negative emotions;<br>Nursing service satisfaction | ① The scores of thoracic surgery-related disease knowledge, nursing points, rehabilitation knowledge, and basic nursing of health knowledge awareness in the observation group were higher than those in the control group;<br>② The SAS and SDS scores in the observation group were lower than those in the control group |

CMFS: Child Medical Fear Scale
SAA: Self-Rating Anxiety Scale
SDS: Self-Rating Depression Scale

In many quality improvement models, patient satisfaction is traditionally measured unidimensional, which suggests that the more the quality level increases, the more satisfaction is attained (Huiskonen & Pirttilä, 1998). Unfortunately, this linear association between healthcare satisfaction and quality is inaccurate and disregards patients’ and users’ emotions and perceptions (Chang & Chang, 2013).

Additionally, patient satisfaction reflects a complex mixture of needs, expectations, and healthcare experiences, which is difficult to measure (Smith, 1992). But Kano Model is based on the two-factor theory, which assumes patients’ satisfaction was not linearly correlated to service functionality.

Furthermore, Kano Model can make the intangible attributes of service quality visible, classify patients’ needs, and use the degree of satisfaction of functional needs to influence user satisfaction. Therefore, applying Kano Model can prioritize service development and improvement, providing a broad understanding of customer needs by identifying the factors that have the greatest impact on satisfaction. With the depth and purpose of the research, the integration of the Kano Model and other quality improvement models occurred. Based on the review outcome, the most common combination is the Kano Model merges with the SERVQUAL Model. The SERVQUAL Model is usually used in public-private hospitals and different populations (Parasuraman et al., 1985a), which calculates the gap between patient expectations and perceptions to enhance healthcare service quality and increase patient satisfaction (Materla, Cudney, & Antony, 2019). The aforementioned statement fails to acknowledge the impact of diverse quality attributes on the level of satisfaction experienced. Therefore, combining with the Kano Model not only remedies this issue but also corrects the shortcoming that the Kano Model fails to consider the perception difference between different customers, which can develop a healthcare service management strategy. The QFD Model’s prominent feature is to collect user requirements using a complex and indirect analytic hierarchy method and convert these requirements into applications through a series of images, intuitive graphics and scientific weighted evaluation methods, and then determines the service standard (Ginting et al., 2020). The problem is that it generally assumes linear relationships between customer satisfaction and the fulfillment of customer requirements. But, a linear function cannot express patient satisfaction only, and there will be an optimal satisfaction value (Kirgizov & Kwak, 2022). According to this view, satisfaction and dissatisfaction are not opposites. Instead, they should be seen as a continuum of various equilibrium states. Hence, the Kano-QFD model can make the Kano Model more operable in measured quality attributes, and potentially promising quality attributes that are often ignored can be effectively addressed.

The application of the Kano Model, regardless of the context, has been found to have the advantage of enhancing the quality of healthcare services. In addition to enhancing patient satisfaction rates, the implementation of the Kano Model has been found to have a positive impact on individuals suffering from anxiety or depression (Zhang Qi et al., 2022). Powell (2018) and Zhang (2022) illustrated that empathy predicted lower distress on average and psychological treatment could reduce patients’ negative emotions. Additionally, it was found that psychological treatment had the potential to alleviate negative emotions experienced by patients. The quality attributes of psychological services, particularly the dimension of empathy, can be considered as essentially one-dimensional categories.

### 4.3 Implication for future research

In Table 3, "information platform" belongs to the one-dimensional category, i.e., patients expect to have an intelligent or Internet-related platform and hospital. In fact, this quality attribute is closely related to the rise of Internet+nursing. Under the concept of patient-centeredness, the components of this system can be identified through the Carnot model. Investigating the classification of patient needs and utilizing the degree of satisfaction of the needs to develop the system, which can satisfy the patient’s needs as well as save resources and improve competitiveness. Also, the mobile health app can be developed in the same way. In short, Kano Model enables healthcare providers to listen to patients and maximize patient satisfaction.

#### Limitation

There exist several limitations to this review. Initially, the research was limited to English and Chinese publications as translation services were not accessible. The publication that has the most influence is typically written in English. Secondly, during the phase of evidence selection, certain articles may be excluded from the review if the search term is not explicitly mentioned in the title or abstract. The search, however, produced a significant number of results, suggesting the relevance of the search. Thirdly, the evaluation of the studies’ quality was not conducted. The limited scope of the review compromised its validity. The present study offers a comprehensive overview of the application of the Kano Model in enhancing the quality of healthcare research. Moreover, it adheres to the accepted methodology of scoping review.

## 5 Conclusion

This review aimed to integrate facts regarding the varied applications of the Kano Model and the map of the most important quality criteria in the healthcare industry. Patient satisfaction is one of the most prevalent measures used to assess the quality of healthcare services. Hence, locating a proper improvement strategy to date is a central theme. We found that reliability, efficiency, and assurance dimensions are essential for preserving patient satisfaction, while the tangible and empathy dimensions are hard to make clear. On the other hand, combining the Kano Model with other models compensate for its flaws and improves the credibility of the research. Meanwhile, "Internet +nursing " has played a crucial role in "high-efficiency and low-risk" healthcare delivery(Yang et al., 2021). Consequently, it is imperative for future research to prioritize the enhancement of internet-based information systems in order to effectively meet the needs of healthcare professionals and patients.

### Funding

This work was supported by the Zhejiang Provincial Hospital Association [grant numbers 2022ZHA-YZJ207] and Health Commission of Zhejiang Province [grant numbers 2021RC011].

### Supplemental material

The supplementary file includes supplementary table (Table S1) which illustrates the original quality attributes.

### Data Available

This is a review article, so data availability is not applicable to this article as no new data were created or analyzed in this study.

## Data Availability

All data produced in the present work are contained in the manuscript.

