## Supplement Table 1 for "The application value of Kano Model in quality of healthcare: A scoping review"

| Author, year, country | Quality attributes | | | | |
| --- | --- | --- | --- | --- | --- |
|  | M | O | A | I | R |
| Wang Shu-yi et al. 2005/China | tangible; professional skill; reasonable guidance; Frequent rounds; therapeutic effect | transportation; service attitudes; patient engagement; Fast process handling | Health education; regular follow-up; | Comfortable signal room; Advice on life of outpatient; | N.A |
| Lee, Chang, and Chao 2007/Taiwan, China | N.A | Nursing Care; Physician Care; Comfort Cleanliness; Courtesy of admissions; Admitting person answered questions. | N.A | Bill; Nearest to convenience; Speed of admissions process. | N.A |
| Chiou and Cheng 2008/Taiwan, China | Convenience; Cleanliness | Nursing care | Physician care | N.A | N.A |
| Tao Yao et al. 2008/China | Cost bill; safety promised; hospitalized environment | Ancillary services; Nurse patrol; waiting time; professional skills; treatment results; medical facilities | Privacy protection; education lecture; provided food; live facilities; | N.A | N.A |
| Hu et al. 2011/ Taiwan, China | Explains how to take the way directions; Equipment of security; Subject labelling; Professional technology; The doctor is punctual; Quality of the drug; Secret degree of the patient; Expenses rationality. | Environment comfortable; Traffic convenience; Parking convenience; Capacity; Moving line; Employee’s clothing; Correct materials; Modernization degree; Insurance service; Medical ethics; Reputation; Commitment; Medical prescription; Attendants; Waiting time; Follows patients regularly; Has patients’ confidence; Security therapy; Accepts covert payments; Examining the purpose; States detailed degree of the prescription; Medical attitude; Listens to patients’ demands; Care for patients; Patients’ interest in the hospital; The improvement situation. | Meals; Community relations; Contribution to the public activity. | N.A | N.A |
| Kuo et al. 2011/Taiwan,China | N.A | Proper medical equipment to patients; Outpatient dept. crew neatly dressed; Easy to access pamphlets of all services; Outpatient time to my convenience; Patients easy to comment and suggest; Finish service accurately during one appointment; Keeping case history correct and complete; Condition improved after treatment and with no complication; Keeping patients informed when and in what way they will be attended; Voluntarily serving patients; Without ignoring patients’ requests for being busy; Professional medical care services convincing patients; Patients feeling safe and credible to be attended; Kind and polite crew; With sufficient knowledge to answer patients’ questions; Understanding patients’ personal needs individually; Providing convenient outpatient time to patient; With accessibility facilities; Well protecting personal privacy; Crew understand patients’ various needs individually | Providing fast services to solve patients’ problems | Beautiful and comfortable outpatient dept. facilities | N.A |
| Cordero-Ampuero et al., 2012/ | N.A | Causes no long-term adverse effects | Achieve symptoms relief allowing daily life activities; Achieve total disappearance of the symptoms; The possibility of resuming activities that are currently not possible due to the disease; Achieve rapid symptom relief; Achieve lasting symptom relief; Once-a-day dosing; Not an injection; Causes no discomfort upon administration; Availability | Advertised in media; It is a new Drug | Recommended by friend or relative |
| Xiong Wei et al. 2012/China | Medical treatment environment; clear sign; medical guideline service; profession skills; | Convenient transportation; waiting time; service attitude; explanation of illness; privacy protection; online appointment; informed consent; | Encouragement of spirit | Health education | N.A |
| ZHANG Zhuo-yi et al. 2013/China | service attitudes; professional skills; communication; hospital environment; nursing care; convenient; patient’s privacy and safety; health education | regular follow-up; reasonable hospitalization expenses; Inform the inspection result in time | A separate bathroom; Provide health advice; waiting time | provide VIP service; Microwave oven provided | No fees for duplicate inspection |
| Su Yin 2013/China | Comfortable environment; clear signs; Guidelines for active consultation after examination; Patiently answer patients' questions; Communicate with children amiable and easy to understand the basic attributes; Children with special needs shall be accompanied by special persons; Wheelchair services for children with behavioral difficulties; Patient comfort for invasive operations; High accuracy of nurse triage; Good infusion and blood drawing technique | Maternal and child room; Service attitude | Convenient facilities for children; | Color wall; cartoon; activity facilities for children | N.A |
| Chou et al. 2015/Taiwan, China | N.A | Employees uniform are clean, nice, and neat; Clean, adequate supplies, and well maintained for every rooms; Meals served are delicious; Employees are helpful, careful, and friendly; No discrimination to the patients | appropriate employees response, medical treatment and doctor visiting are well scheduled, available and adequate patient family visiting time, the employees solve the elderly problem sincerely, all equipment work properly | all patient activities are well scheduled | N.A |
| LI Teng-hun et al. 2015/China | Sanitation of the medical environment; service attitudes; | Timeliness of disease diagnosis; Convenience of taking medicine; Medical expenses; Interrogation time | Clinic signage; description of the condition, medication use, and course of treatment | Language used; privacy protection | N.A |
| Zhai et al. 2015/China | Professional skills | Setting up clinics for midwives; Have perfect advanced medical equipment; Have a brief introduction materials; The environment of the clinic is warm; Patiently answer the patient's questions; Caring for patients and listening to their needs; Discuss guidance programs with pregnant women; Timely admission to hospital; Quickly complete admission procedures and be admitted to the ward | The temperature is suitable; There are comfortable private rooms; Providing a variety of educational formats; Able to make house calls at night and holidays; Regular follow-up and active reminder for puerpera; To provide comprehensive physical and social education on childbirth; Advice on the life of the；Providing health care knowledge during the prenatal and midterm periods; Detailed instruction; Advice on the life of the puerperia; To provide specific guidance on hospitalization matters; Continuous whole-course team service; Improved quality of delivery; Reduce the incidence of complications; | N.A | N.A |
| Nie Shijun 2016/China | Clean and comfortable environment; clear and concise emergency signs; Respect the patient's right to informed consent; health education; provide list of charge | Professional skills; privacy protection; Expedite admission procedures; Friendly attitude; response patients on time; Actively participate in rescue work | Service attitudes; Monitoring equipment | N.A | N.A |
| Li Zhen 2017/China | Patiently answer the children's family questions; Communicate with children kindly and easy to understand; Children with special needs shall be accompanied by special persons; Provide basic attributes of wheelchair services for children with behavioral difficulties; Patient comfort for invasive operations; High accuracy of nurse triage; Good infusion and blood drawing technique; The comfortable waiting environment; | A feeding room is provided; The triage nurse smiles and serves actively | Convenient facilities for children | The walls are brightly colored; Facilities for children's activities; Play cartoons | N.A |
| Patel and Bhatt 2017/India | Reliability; Tangibility | Assurance; Responsiveness; Empathy | Willingness of hospital personnel to help patients; Blood bank within the premises | N.A | N.A |
| Tian Yuan et al. 2017/China | Professional skill; Auxiliary tools; handle problem immediately; No complications; | Service attitudes; being patient and satisfy reasonable requirements; Explain the precautions; Provide rehabilitation guidance; | health education; Provide convenience items; regular follow-up; | Establish nurse-patient communication cards | N.A |
| Gomez Martin et al. 2019/Spanish | Free television; Television automatic turn off at midnight | N.A | Individual room; Information about the exact time for the dressing change; More additional staff for emergencies | Psychological treatment for relatives Cleaner rooms Waiting time at the office shorter than 30min Provision of telephone consultations | N.A |
| Gong et al. 2019/China | The comfortable environment; Clear sign; Reasonably arrange the physical examination of critically ill children; Timely detection of disease changes; High triage accuracy; Skilled in professional technology; Patiently explain the problem; Be amiable when communicating; Explain the precautions in detail | Maternal and child room; Effective sedation measures should be taken before examination. Active smile service; Frequent ward rounds; Placate the patient during puncture; Special person to accompany special patients; Proactive health education | Broadcasting health education content; Set up a special platform to acquire relevant knowledge; brightly colored wards | N.A | N.A |
| Rozaq et al. 2019/Indonesia | N.A | Skill; innovation; physically accessible; completeness; ability to provide operations and the promised service on time; availability of all kind of services at the hospital; infrastructure; courtesy; standard | N.A | Experience; caring; manner; communication; timely; willingness; automatic; cost; compensation; | N.A |
| Howsawi et al. 2020/Saudi Arabia | N.A | Distance from home to the PHC; A parking lot; Working hours of the clinic; Cleanliness of the PHC; Token display system; Friendliness and politeness of the clinic receptionist; Friendliness and politeness of the nurses and laboratory staff; Care and attention of the doctor; Quick response by the doctors; Quick response by nurses and laboratory staff; Advanced laboratory services such as cultures; Electronic referral system; Examination explained by doctors; Get informed about the medical condition | Display of educational films in the waiting room; Unified electronic medical record; Advanced radiology services such as MRI | Minor operating room | N.A |
| Hun Chen et al. 2020/China | Environmental safety; Change bed sheets; clean restroom; The charges are clear; professional skills; introduction inpatient environment; Detailed information about the condition and treatment options; Solicit opinions on special examinations or expensive drugs or self-paid drugs; Regular ward round | Quiet ward; Registration is simple and convenient; High efficiency of hospitalization procedures; Good medical ethics; Be respected; Treatment is effective; Check that the waiting time is reasonable; Be kind and polite; Health education; Dietary guidance; Listen to the patient's description; | The ward provides greenery; A nutritious diet; The inspection results are easy to query; Moral encouragement | N.A | N.A |
| He et al. 2020/China | Friendly attitude; Respect patients; Physical examination; Ask the history of drug allergy, ask women of childbearing age whether they are pregnant or lactating; According to the condition of the examination sheet; Doctors explain and communicate the conditions and risks; Prescribing medication according to the condition; Safety information of medication | The diagnosis is correct; Treatment is effective; The cost is within the estimated range | Strong ability to solve difficult problems; Explain the etiologic, pathogenesis, pathophysiology and prevention of diseases to patients; Health education; | N.A | N.A |
| Tejaswi Materla and Cudney 2020/the USA | medical staff is appropriately qualified to provide care | medical staff includes you in the decision-making process; the design of a walk-in clinic facility is patient friendly; the rooms allow for personal privacy; medical staff provides written communication of how your treatment will be delivered; patient check-in process is easy; the staff keeps you informed about the delays; medical staff provides correct care on the first time; walk-in clinic personnel have good communication among them to assure effective treatment; walk-in clinic personnel provide clear instructions about follow up care; medical staff are sympathetic and reassurin; the medical staff provide adequate information about your illness and treatments; walk-in clinic personnel provide complete information of the prescribed medications; walk-in clinic personnel are friendly; medical staff understands your needs and requirements; feel confident about the care provided by the medical staff | qualified medical staff were available within ten minutes; after-hours care is provided by a walk-in clinic | the walk-in clinic personnel accommodate your religious restrictions when conducting your medical care; walk-in clinic personnel accommodate your cultural restrictions when conducting your medical care | N.A |
| Barrios-Ipenza et al. 2021/Peru | making a medical appointment; the level of bureaucracy; resolution of complaints | health personnel; Non-Health Personnel; Facilities, Equipment, and Tangibles; Efficiency | N.A | N.A | side effects of medicines |
| Ke, Chen, and Gou 2021/China | Working attitude of guidance desk; The service of registered fee collection staff; The therapeutic effect is expected; Consultation on doctor's choice of treatment; Detailed bills issued by the hospital at discharge; Outpatient Environmental Hygiene; Canteen food satisfaction; Follow up platform to query personal medical information; The follow-up platform provides communication services with doctors and nurses | The service attitude of the doctors you contact; The service attitude of the nurses you contacted; The service of ultrasound department staff; Doctor's explanation of the condition and medication; Time consuming for registration, pricing and payment | Services of laboratory staff; Service of radiology staff; Outpatient medicine timely, convenient; Clarity of department floor layout; Follow up rehabilitation guidance; The questions were answered during the follow-up | Service of pharmacy staff | N.A |
| Lacerda et al. 2021/ Brazil | N.A | Tangibles; Reliability; Responsiveness; Assurance; Empathy | N.A | N.A | N.A |
| Mehrabian, Gilani, and Almaee 2021/ Iran | N.A | observance of sanitary by the service staff; Proper arrangement of hospital equipment (e.g., beds, wardrobes) and rooms; Fixing possible problems and shortcomings by staff in a timely manner; Feeling emotionally comfortable after being in the hospital; Proper fulfilment of Health Transformation Plan for patients; Use of signs and warnings to increase patient information in the hospital; Use of modern equipment in the hospital; Employees’ interest in providing services to patients such as wheelchairs and european toilets; Creating trust, physical and financial security in the patient by the hospital staff; Suitable food for the patient; Having enough staff to provide timely services | Neat and tidy appearance of all hospital staff; Performing services on time; Appropriate working hours for staff in the hospital to provide services; Ease of access and contact with the hospital and no busy phone lines; Employees ‘ability and interest in responding to patients’ complaints; Employing specialized staff; Providing clear and complete information to the patient at the time of admission, and information on possible cost rates; Speed of service delivery in the hospital; Informing patients about the end date of hospitalization; Appropriate location of the hospital ;cleanliness of the whole hospital; Appropriate location and distribution of hospitals in the city; Demanding the best benefits for patients from the management and staff in hospital | N.A | Possibility of filling the admission online early before going to the hospital |
| Ferreira et al. 2021/ Portugal | Facilities; food; doctor; Volunteering; Discharge process | Efficiency; Concern with the child; Cleanliness and hygiene | Capacity of providing useful information; Visits; nurse; Auxiliary staff; Administrative staff; Diagnosis and treatments | N.A | N.A |
| Wang Huaiyan et al. 2021/China | Hospital environment; service attitude; establish health card | Maternal and child room; Psychological care; Health knowledge and education; | Convenient equipment; professional skills; | Set up a complaint mailbox | N.A |
| Zhang Qunxiang et al. 2021/China | Emergency response capacity; To protect privacy | Convenient transportation; Online registration; convenient process; Diagnosis and treatment effect; Reasonable charges; Professional personnel; Communication; Waiting time for treatment; Respect for patients; Rational drug use; | Advanced equipment; Internet+ diagnosis; Service attitude; To accept complaints | N.A | N.A |
| de Vasconcelos et al. 2022/Brazil | Care Nursing; Managers (Involvement Quality); Location; Business Hours; Readiness | Service at Reception; Consultations Marking; Medical Care; Installations; Internal Layout; Service Reception ; Personalization; Modern Equipment; Credibility; Service at Reception; Medicines; Readiness; Knowledge | N.A | N.A | N.A |
| He Jian-qun et al. 2022/China | Clean environment; Signage Guidelines; completed equipment; exact result; reasonable costs; insurance consultant; understandable expression; privacy protection | Online booking; convenience transportation; clean flow; waiting time; proficient skill; reasonable advice; clear bills; | Guiding service; consultant; | Medical guidance | N.A |
| Chen et al.2022/ China | Respect patient’s right; service ability and level; hospital environment; | Convenience of medical treatment; outpatient expenditure; physical examination; communication | N.A | N.A | N.A |
| Luming Li et al. 2022/China | Professional skill; Nurse response speed; privacy protection; communication; communicate with the patient before using the medication; discharge expenses | Application of new medical technologies; clean ward; Admission procedures procedure; service attributes; Explanation of Precautions; quality of food | Ward night environment; Medical examination time  Ward rounds frequently; health education; medical cost | Road signs and instructions | N.A |
| XIONG et al. 2022/China | Clean and comfortable environment; precious signs; convenient stopping station; | Service attitude; understand patients’ needs; High reliability self-service system | Leisure and entertainment facilities; online review platform; full functional and easy operation of self-service system; Voice interactive intelligent robot; | N.A | N.A |
| Cudney et al., 2023/ the USA | N.A | Correct care provided the first time; Staff listens to patients about care goals; Necessary supplies are provided for caring for the wound; Clear instructions provided on follow up care; Staff is appropriately qualified to provide care; Staff understands the impact of the wound on patient's independence; Personnel are trained in wound care; Rooms allow for privacy; Adequate information provided about treatment;  Informed of delays during visit; Check-in process is easy; Confidence in the care provided; Complete information provided on prescribed medications; Communication amongst staff for effective treatment; Written communication provided on treatment plan; Included in decision making process of healthcare delivery. | After-hours care available | N.A | N.A |
| Zhu et al., 2023/ China | Effective sedation prior to examination; Effective sedation during seizures; Timely notice of changes in the child's condition; No nursing complications occur; Patience in explaining family issues; Kindness in communication with children; Able to provide detailed answers to questions about convulsions; Able to answer in detail the role of tests, treatments and medication; Able to explain in detail the side effects of anticonvulsant therapy; Ability to answer family questions in detail and persuasively; Able to keep the family informed of changes in the child's condition | Proactive service with a smile; Proactive guidance upon inspection completion; Nurse with gentle words; Bedside examination of comatose children whenever possible; Able to pacify children and prevent them from rejecting EEG monitoring(*) equipment; Involve family members in the care of children as much as possible; Setting up a convulsion recovery hotline; Precautions to be taken prior to examination; Instruct families in proper life care; Help families understand the child's behavioural responses. | Patiently calming the child during the operation; Play cartoons; Play health education content; There are dedicated platforms for accessing relevant knowledge | N.A | N.A |
| Wang et al., 2023/ China | Listen to me talking about my spiritual concerns; Help me to enjoy quiet times or space. | Teach me about ways to draw or write about my spirituality; Listen to the stories of my life; Offer to talk with me about meditation; Ask me about what gives my life meaning; Ask me about my spiritual beliefs. | Listen to me talking about my spiritual strength; Help me to think about my dreams; Bring me humorous things, e.g., share a joke. | Ask me about religious practices; Help me, if I needed, with my religious practices. | N.A |
| 姚瑶 et al., 2023/China | Provide adequate maternal and fetal assessment and monitoring; Regular checkups; Regular weight monitoring; Reduce the use of expensive consumables during hospitalization; Postpartum uterine contractions are good; Postpartum urination; Less postpartum bleeding; No vaginal hematoma after delivery; Good maternal and infant outcomes | Self-service archiving; Provide automatic pregnancy test reminder; Provide remote fetal heart monitoring during pregnancy; Provide comprehensive health education; Provide convenient information exchange platform; Short waiting time for prenatal check-up; Midwives trained in systematic midwifery skills; shorter hospital stay | Perineum flexibility training instruction; Free position during labor and delivery; Adequate protection of the perineum during labour and delivery; Midwives have good birth injury control skills; Midwives help ease childbirth pain; Midwives control delivery speed; Postpartum recovery | N.A | N.A |
| 孙卫格 et al., 2023/ China | Preoperative preparation information; Dietary guidance | Disease-related information; Psychological intervention; Whole process management platform; Surgical information; Postoperative follow-up; Follow-up procedure information; Take out stitches dressing; Drug usage | Online consulting platform | Drug storage information | N.A |

**Table S1** Summary of original quality attributes
